# Haemolysis detection in microRNA-seq from clinical plasma samples

**DOI:** 10.1101/2022.03.27.22273016

**Authors:** Melanie D. Smith, Shalem Y. Leemaqz, Tanja Jankovic-Karasoulos, Dale McAninch, Dylan McCullough, James Breen, Claire T. Roberts, Katherine A. Pillman

## Abstract

The abundance of cell-free microRNA (miRNA) has been measured in many body fluids, including blood plasma, which has been proposed as a source with novel, minimally invasive biomarker potential for several diseases. Despite improvements in quantification methods for plasma miRNAs, there is no consensus on optimal reference miRNAs or to what extent haemolysis may affect plasma miRNA content. Here we propose a new method for the detection of haemolysis in miRNA high-throughput sequencing (HTS) data from libraries prepared using human plasma. To establish a miRNA haemolysis signature in plasma we first identified differentially expressed miRNAs between samples with known haemolysis status and selected miRNA with statistically significant higher abundance in our haemolysed group. Given there may be both technical and biological reasons for differential abundance of signature miRNA, and to ensure the method developed here was relevant outside of our specific context, that is women of reproductive age, we tested for significant differences between pregnant and non-pregnant groups. Here we report a novel 20 miRNA signature (miR-106b-3p, miR-140-3p, miR-142-5p, miR-532-5p, miR-17-5p, miR-19b-3p, miR-30c-5p, miR-324-5p, miR-192-5p, miR-660-5p, miR-186-5p, miR-425-5p, miR-25-3p, miR-363-3p, miR-183-5p, miR-451a, miR-182-5p, miR-191-5p, miR-194-5p, miR-20b-5p) that can be used to identify the presence of haemolysis, *in silico*, in high throughput miRNA sequencing data. Given the potential for haemolysis contamination, we recommend that assay for haemolysis detection become standard pre-analytical practice and provide here a simple method for haemolysis detection.

## Introduction

MicroRNAs represent a class of short, ∼22 nt single stranded non-coding RNA transcripts found in the cytoplasm of most cells that act directly as post transcriptional regulators of gene expression (1,2) and also coordinate extensive indirect transcriptional responses (3). In their canonical action, miRNAs mediate the expression of specific messenger RNA (mRNA) targets by binding to the 3’-untranslated region (UTR) of transcripts by either repressing translation or marking them for degradation (4).

In the canonical miRNA pathway, target specificity requires exact nucleotide sequence complementarity between the miRNA ‘seed’ region (the first 2-7 bases at the 5’ end of the mature miRNA transcript) and the 3’-UTR of the mRNA. Importantly, miRNAs demonstrate tissue, temporal and spatial expression specificity and are known regulators of development, with most mammalian mRNAs harbouring conserved targets of one or many miRNAs (1,2,5).

miRNA expression is both temporally and spatially tissue-specific, with transcripts identified beyond the cells in which they were synthesised, in various body fluids including urine, saliva and blood plasma (6). Circulating cell-free miRNAs identified in plasma are packaged in micro vesicles such as exosomes (7,8) or bound to protein complexes such as argonaute 2 (Ago2), nucleophosmin 1 (NPM 1) and high density lipoprotein (HDL) (9–11), making them exceptionally stable (6). This stability, coupled with their minimally invasive accessibility, has suggested circulating cell-free miRNAs as an important resource for the identification of novel biomarkers.

Whilst much progress has been made in the search for novel miRNA biomarkers of disease processes (12–14), outcomes of this research approach are often inconsistent or even contradictory (6). There are many reasons for this, including variations in enrichment, extraction and quantification methods, variation between individuals, lack of consensus regarding optimal reference miRNA for normalisation and the difficulty in quantifying both the amount and quality of RNA transcripts from blood plasma samples (15,16). An important but often overlooked factor is the potential for sample haemolysis during blood collection or sample preparation which results in miRNA from lysed RBCs being spilled into and retained within the plasma sample to be assayed (15).

The issue of haemolysis altering the miRNA content of plasma and the potential for confounding biomarker discovery has been reported previously (15,17,18). Using RT-qPCR Kirschner and colleagues (15) showed that contamination of plasma samples with the miRNA content of RBCs changed the abundance of both miR-16 and miR-451a. This, in turn, altered the relative abundance of potential biomarkers for mesothelioma and coronary artery disease including miR-92a and miR-15. Using the same technique, Pritchard *et al*. (18) demonstrated in plasma that 46 of the 79 circulating miRNA cancer biomarkers were highly expressed in more than one blood cell type, noting that the effects of sample specific blood cell counts and haemolysis can alter the miRNA biomarker levels in a single patient sample up to 50-fold. As a result, the authors emphasised caution in classifying blood cell associated miRNAs as biomarkers given the possible alternate interpretation.

Haemolysis is associated with either blood collection or RNA extraction and sample preparation. Thus, despite differences between the quantification methods, high throughput sequencing data used in our study is equally susceptible to the confounding effects of sample haemolysis on miRNA abundance levels in plasma is RT-qPCR. There are currently two gold standard approaches in the assessment of haemolysis in plasma: 1. Delta quantification cycle (ΔCq), where expression levels of a known blood cell associated miRNA (miR-451a) and a control miRNA (miR-23a) are determined based on the difference between the two raw Cq values and 2. Spectrophotometry, where absorbance is measured at 414 nm with the use of a spectrophotometer. In the case of ΔCq assessment, miR-451a is known to vary and miR-23a is known to be invariant in plasma affected by haemolysis (15,16). Using spectrophotometry, haemolysis is quantified by assessing the presence of cell free haemoglobin by measuring the absorbance at 414 nm, the absorbance maximum of free haemoglobin (19,20). Both methods require access to sufficient amounts of the original plasma sample and the laboratory equipment required to perform the assays. Free access to a web-tool that can perform *in silico* assessment of RBC contamination in human plasma would be of exceptional value to the research community.

Whilst it is well established that haemolysis frequently occurs during extraction or processing of blood samples, the assessment of RBC contamination is rarely mentioned in publications. It is even more rare that the results of any such testing are present in the metadata assigned to publicly available sequencing data. There is currently no publicly available tool for analysis of haemolysis without access to the physical plasma specimen. Although the theory underlying identification of haemolysis in plasma is relatively straight-forward, surprisingly, to our knowledge this has never before been extrapolated into a data-only, *in silico* approach. The paucity of haemolysis information in the context of publicly available datasets, combined with the lack of tools to identify affected datasets after the fact, substantially limits the utility of this data resource and reproducibility of research findings. Further, it increases the risk that results obtained may unwittingly represent blood-cell based phenomena rather than signatures of the pathology of interest.

In this study, we assessed miRNA abundance in HTS data from libraries prepared using human plasma from pregnant and non-pregnant women of reproductive age. Using a set of samples with confirmed haemolysis (ΔCq (miR-23a-miR-451a)), we established a set of 20 miRNAs differentially abundant between plasma from samples with and without substantiated haemolysis. Using the expression values of these 20 miRNAs as a ‘signature’ of haemolysis, we calculated the difference between the mean normalised expression levels of these miRNAs compared to those of all other miRNAs (as a ‘background’ set). This produced a quantitative metric which represents the strength of the evidence of haemolysis in an individual sample. When this metric is interpreted in the context of other samples, it can be used to identify sample(s) that display substantial evidence of haemolysis. The researcher may consider discarding these samples from further analyses or using caution in their interpretation. We consulted the EMBL-EBI Expression Atlas (ebi.ac.uk) to ensure all signature miRNAs are identified in multiple human tissues (male and female) and have no known developmental stage association. For ease of application, we have developed this method into a web based Shiny/R application, DraculR (a tool that allows a user to upload and assess haemolysis in high-throughput plasma miRNA-seq data), for use by the research community (DraculR: A web-based application for *in silico* haemolysis detection in high throughput small RNA sequencing data).

## Results

### High throughput sequencing

Illumina NextSeq 75 bp single-end read sequencing was performed on miRNA libraries from 154 plasma samples taken from 24 non-pregnant and 130 pregnant women aged 16 to 46 years (Qiagen, Hilden, Germany). Prior to sequencing, RT-qPCR was used to analyse ΔCq (miR-23a-miR-451a), where the ratio of miR-23a to miR-451a (or ΔCq (miR-23a-miR-451a) ≥7) correlates with the degree of haemolysis. We identified 14 plasma samples with a ΔCq of 7 or above (Supplementary Table 1). An average of ∼2.9 million reads were sequenced per sample (range ∼0.25-18.6 million reads). Thirty-one libraries with < 1 million reads were considered to be unreliable due to low sequencing output and were removed from further analyses. There was no difference in the proportion of haemolysed and non-haemolysed data in the exclusion of samples due to low library size (Fisher’s exact test P-value = 0.7). Sequence alignment was performed using BWA (21) to the human genome (version GRCh38) and miRNA read counts were generated by mapping to human miRBase v22 (22,23) identifying 1,133 mature miRNAs.

To analyse the effects of haemolysis on miRNA expression data from next generation sequencing, we first determined the number of unique mature miRNAs identified in each of our samples and analysed the data relative to read depth. Using an analysis of variance (ANOVA) we identified a significant difference between the haemolysed and non-haemolysed samples (P<0.05), with haemolysis being frequently associated with fewer mature miRNA species detected at a given read depth (Figure 1).

**Figure 1.**
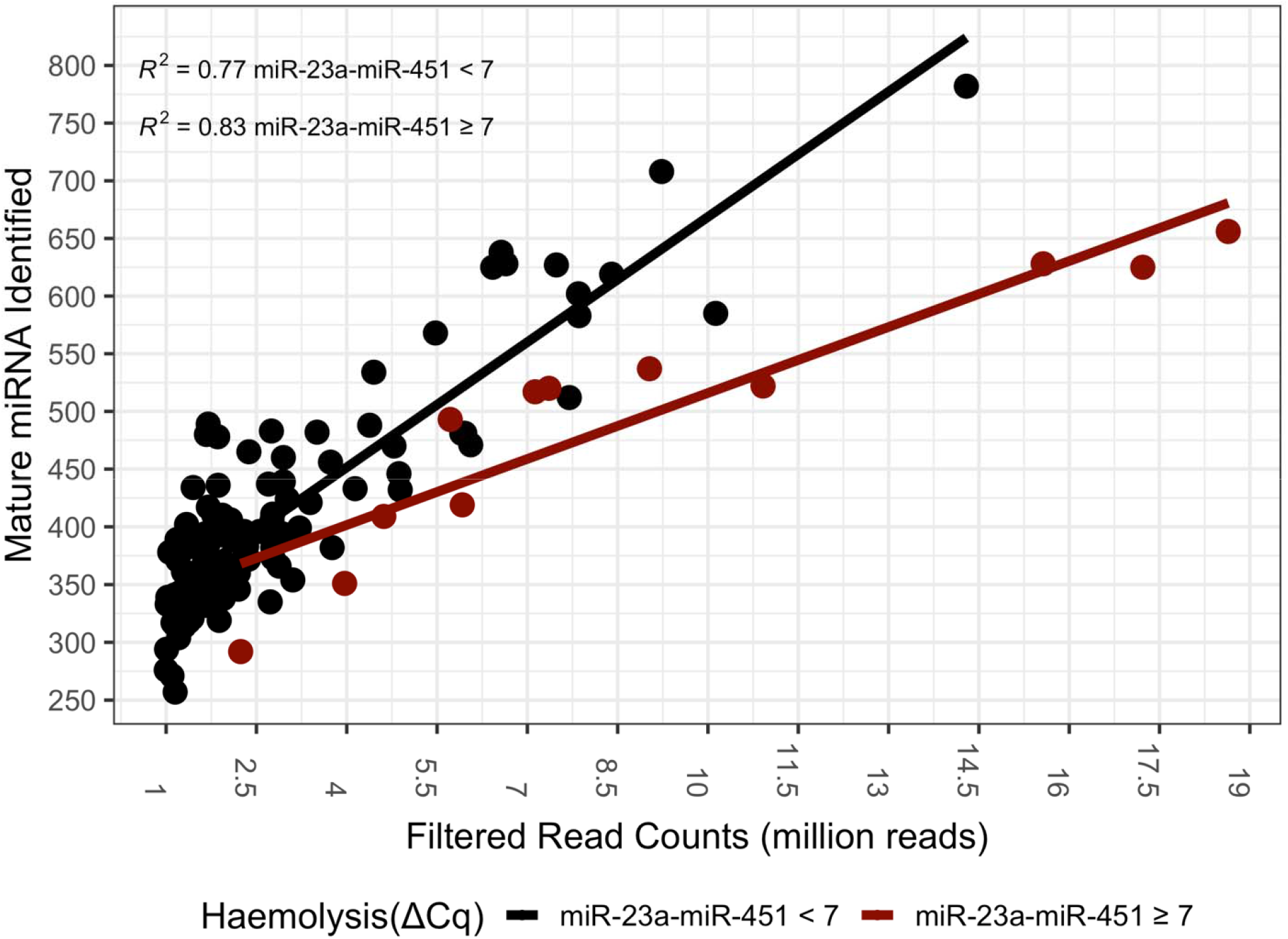
The number of mature miRNA species identified in an individual sample increases with read depth for both haemolysed and non-haemolysed samples. However, the number of mature miRNA species identified for a given read depth is significantly lower (ANOVA, P-value = 1.68 × 10^−9^) in samples affected by haemolysis (maroon) when compared to a non-haemolysed sample (black) of equal read depth.

### microRNA haemolysis signature set

To ensure that miRNAs identified here were representative of those found in a broad set of plasma samples, we first filtered to discard miRNAs of low abundance. After filtering, 189 highly abundant miRNA remained. Differential expression analysis comparing miRNA read counts identified 138 miRNAs with a higher abundance in haemolysed compared to non-haemolysed samples (statistically significant differentially expressed miRNA, false discovery rate (FDR) < 0.05, with a log_2_ fold change (log_2_FC) > 0) (Supplementary Figure 1; Supplementary Figure 2a & b). We further ranked the differentially expressed miRNAs based on log_2_FC, FDR and abundance levels and subset the list such that only miRNAs which had a log_2_FC > 0.9 and were in the top 60 percent of each of the FDR and abundance rank criteria remained. This resulted in a high confidence set of 20 miRNA indicative of a haemolysis signature (Table 1).

**Table 1.** 20 miRNAs with a general-use plasma haemolysis signature set. To remove confounding effects within our pregnancy-specific dataset, we identified a subset of 10 abundant miRNA which are invariant with respect to pregnancy.

| miRNA | Log<sub>2</sub>FC | Average Expression (log<sub>2</sub> CPM) | Adjusted P-value | Pregnancy Assoc. |
| --- | --- | --- | --- | --- |
| miR-106b-3p | 1.589 | 8.731 | 8.61 x 10<sup>-15</sup> | no |
| miR-140-3p | 1.073 | 10.098 | 2.75 x 10<sup>-13</sup> | no |
| miR-142-5p | 0.962 | 10.651 | 4.96 x 10<sup>-12</sup> | no |
| miR-532-5p | 1.288 | 7.237 | 4.96 x 10<sup>-12</sup> | no |
| miR-17-5p | 0.952 | 7.892 | 7.84 x 10<sup>-12</sup> | no |
| miR-19b-3p | 1.128 | 8.696 | 1.93 x 10<sup>-09</sup> | no |
| miR-30c-5p | 0.95 | 7.325 | 2.48 x 10<sup>-09</sup> | no |
| miR-324-5p | 1.304 | 7.186 | 2.50 x 10<sup>-09</sup> | no |
| miR-192-5p | 0.941 | 8.944 | 1.37 x 10<sup>-08</sup> | no |
| miR-660-5p | 1.305 | 7.62 | 3.45 x 10<sup>-10</sup> | no |
| miR-186-5p | 1.228 | 8.052 | 2.75 x 10<sup>-13</sup> | yes |
| miR-425-5p | 1.282 | 11.246 | 4.96 x 10<sup>-12</sup> | yes |
| miR-25-3p | 1.212 | 12.939 | 1.26 x 10<sup>-11</sup> | yes |
| miR-363-3p | 1.237 | 7.882 | 4.52 x 10<sup>-11</sup> | yes |
| miR-183-5p | 1.55 | 9.382 | 9.34 x 10<sup>-11</sup> | yes |
| miR-451a | 1.372 | 13.002 | 3.65 x 10<sup>-10</sup> | yes |
| miR-182-5p | 1.341 | 10.585 | 2.48 x 10<sup>-09</sup> | yes |
| miR-191-5p | 0.929 | 11.79 | 4.68 x 10<sup>-09</sup> | yes |
| miR-194-5p | 0.937 | 7.679 | 1.85 x 10<sup>-08</sup> | yes |
| miR-20b-5p | 0.932 | 7.43 | 1.96 x 10<sup>-08</sup> | yes |

For *in silico* assay of haemolysis in our data, we further removed miRNAs associated with pregnancy to avoid confounding miRNA associated with haemolysis with those associated with pregnancy. Differential expression analysis of miRNA read counts from pregnancy and non-pregnancy samples identified 127 miRNAs (FDR < 0.05) that were significantly differentially expressed between the groups (Supplementary Figure 3a & b). Strikingly, one of our first observations highlighted the importance of including haemolysis analysis as an adjunct in our study: miR-451a, which is the sole haemolysis signature miRNA used in the current ΔCq (miR-23a-miR-451a) gold standard method for haemolysis detection was discovered to be highly correlated with pregnancy status, indicating a strong confounding factor in pregnancy studies when haemolysis levels are estimated using RT-qPCR alone. Accordingly, miR-451a was removed from calculations hereafter along with 9 other miRNAs that were differentially expressed between the pregnant and non-pregnant groups from the core set of haemolysis signature miRNA. This resulted in 10 miRNAs remaining for evaluation of haemolysis levels.

Incorporating concepts from previous RT-qPCR analyses of haemolysis, we established a new measure of the inclusion of RBC associated miRNA in human plasma. After establishing the 20-miRNA signature associated with RBC content inclusion, we determined the geometric mean of the distribution of miRNA read counts as an appropriate measure of abundance and summary statistic. Using this summary statistic, our method calculates a ‘Haemolysis Metric’, defined as the difference between the geometric means of the normalised abundance levels of the haemolysis miRNA signature set compared to that of all other miRNAs (the ‘background’ set). Note that in a case-control study, to reduce the risk of confounding the Haemolysis Metric with experimental variables, the signature set should be reduced to exclude any miRNA known to be differentially expressed between groups. In this case, the geometric mean of the reduced signature set will be calculated, as defined in (1).

Let

*Zx*. be the miRNA Reduced signature set (log_2_ CPM counts)

and *Zy*. be the Background miRNA set (log_2_ CPM counts)

where *x* = 1,2,3,…, *p*_1_ with *p*_1_ = the number of miRNA in Reduced signature set

and *y* = 1,2,3,…, *p*_2_ where *p*_2_ = the number of miRNA in Background

and *i* = 1,2,3,…, *n* where *n* = the sample size after filtering

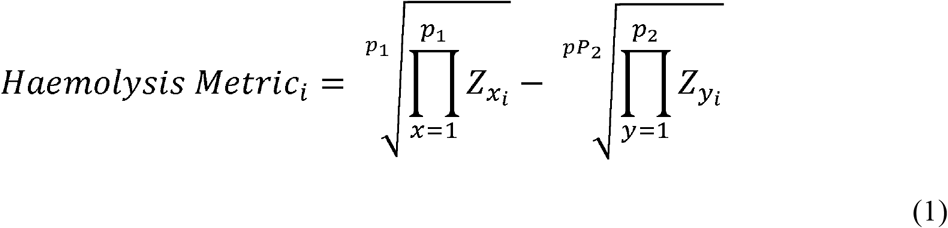

Prior to establishing a threshold for the new Haemolysis Metric we measured the linear dependence between the new Haemolysis Metric and the ΔCq (miR-23a-miR-451a) metric by performing a Pearson’s correlation. Our results indicated a Pearson’s correlation coefficient of 0.64 (P < 0.0001). With confidence in the correlation, to establish a threshold for the Haemolysis Metric, we compared the results of the ΔCq (miR-23a-miR-451a) and summary statistic methods directly. Briefly, we compared the Haemolysis Metric to the ΔCq (miR-23a-miR-451a) results for matched samples and established a cut off criterion for inclusion into the Clear (no haemolysis detected) and Caution (haemolysis detected) groups (Figure 2a). We chose a threshold of ≥ 1.9 for the assignment of “Caution” to individual samples based on the minimum summary statistic difference of samples assayed using the ΔCq (miR-23a-miR-451a) metric of ≥ 7 (Figure 2a) and the minimal overlap between the distribution of the Haemolysis Metric in haemolysed compared to non-haemolysed samples (Figure 2b). Where a sample is assigned “Caution”, researchers are advised to consider removing the sample, or to continue with caution. Given the correlation of the two metrics is imperfect and the arbitrary nature of choosing any cut-off, samples with a Haemolysis Metric close to the 1.9 cut-off may be interrogated further prior to any decision to retain or remove. Of the 121 samples assayed, 25 samples met the criteria for Caution. Of these, 12 were previously determined as haemolysed or borderline using the ΔCq (miR-23a-miR-451a) assay. We found that all samples identified as ΔCq ≥ 7 (Figure 2a, scarlet) are above the criteria for the Haemolysis Metric (Figure 2a, horizontal grey bar; threshold ≥ 1.9). Further, we identified 13 samples with a Haemolysis Metric ≥ 1.9 not included in the ΔCq (miR-23a-miR-451a) criteria.

**Figure 2:**
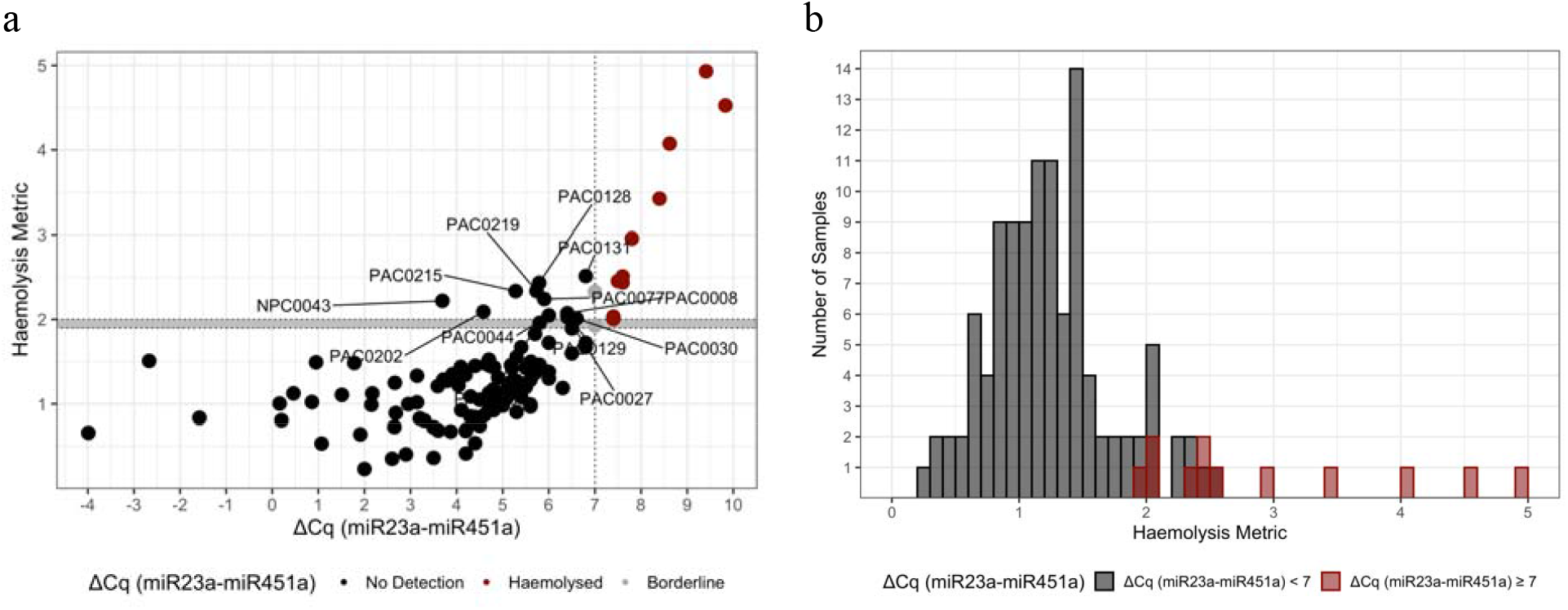
(a) A comparison of the derived Haemolysis Metric and the ΔCq measure of haemolysis shows a clear correlation. We identified 13 samples (named) that we suggest should be discarded or used with caution in further analysis. (b) Histogram of Haemolysis Metric values from the 121 samples in our experiment, coloured according to their ΔCq (miR-23a-miR-451a) classification indicate a minimum Haemolysis Metric of ≥ 1.9 for samples previously identified as haemolysed.

## Discussion

Through an analysis of differential miRNA expression in samples whose haemolysis levels were known, we identified a novel 20 miRNA signature indicative of haemolysis. Given our hypothesis that plasma samples contaminated with RBC content would contain proportionally higher levels of many RBC-associated miRNA, not just miR-451a, we established a method using a group of background miRNAs as a reference. Accordingly, as a group, signature miRNAs (microRNAs which are abundant in red blood cells) are shown to be more highly abundant in samples contaminated with RBCs. The degree of this change can be used as a measure of RBC content contamination and quantified by comparing the geometric means of the expressions of RBC signature miRNAs to that of the background set of miRNAs. We further established that where a comparison between conditions is considered, for example in a biomarker discovery experiment, any miRNA known to be associated with the condition for which the biomarker is proposed should be removed to prevent confounding between the condition of interest and the quantification of RBC-associated miRNA inclusion.

Our experimental results demonstrate that it is possible to identify a haemolysis signature *in silico*, avoiding the effort and expense of lab validation, and in situations where blood plasma samples are exhausted, otherwise unavailable or cost-prohibitive to assay using current gold standard approaches. Given the limited access to physical samples associated with publicly available data, the Haemolysis Metric technique introduced here, provides the basis for the development of a publicly available tool (DraculR: A web-based application for *in silico* haemolysis detection in high throughput small RNA sequencing data).

Among the haemolysis miRNA signature is miR-451a (previously named miR-451), commonly associated with RBC contamination and used in the calculation of ΔCq (miR-23a-miR-451a). However, we removed miR-451a together with 9 other miRNAs from our calculation of distribution difference due to changes in miRNA abundance associated with pregnancy. During pregnancy total blood volume increases, varying between 20% to 100% above pre-pregnancy levels. This change, however, is not uniform across all blood components as plasma volume increases proportionally more than the RBC mass (24). This is an important consideration and highlights the limitation of the current gold standard approach that uses two miRNAs rather than a larger signature set to calculate a measure of contamination. If, as in this example, the abundance of either miRNA used to determine the ΔCq is also affected by the condition or pathology under investigation, the issue is two-fold. Firstly, you may identify miR-451a as being differentially abundant in the pathology of interest and propose its use as a biomarker only to find that it is confounded by haemolysis. Secondly, you may, using the ΔCq calculation, classify samples as haemolysed when the change in miR-451a abundance is more appropriately associated with the pathology of interest. By establishing a larger signature set of miRNAs to detect haemolysis in small RNA sequencing from human plasma we hope to provide a resource to the community. To overcome issues identified in previous studies, the flexibility and redundancy included in our metric buffer against the issue of confounding conditions of interest with the measure of haemolysis.

We found limited overlap between the miRNAs identified as useful for the detection of haemolysis and those previously reported as markers of haemolysis contamination (15,18,25). When comparing with previous research, it is important to note, that our research question differed from that of the above studies, as did our study methodology. The most important technical difference is in the quantification of miRNAs. The expression values used here were taken from a HTS experiment, rather than RT-qPCR used previously. The limitations of RT-qPCR to investigate which miRNAs are affected by haemolysis has been identified previously (26). Given that HTS allows for quantification of all known miRNA species and that RT-qPCR is targeted, our experiment was able to identify differential abundance in miRNAs not quantified in Kirschner *et al*. (15), Pritchard *et al*. (18) or McDonald *et al*. (25). Whilst we identified an overlap in the miRNAs associated with haemolysis in this and previous work, many of these were not included in the final miRNA Haemolysis Metric signature set. These include miR-16, miR-486-5p and miR-92a-3p which were significantly upregulated in the haemolysed group, but not included in the signature set as they failed to pass filtering criteria for log_2_FC and expression level. Secondary to the technical differences introduced by using different miRNA quantification technologies, it is important to note that all plasma samples used here to establish which miRNAs are affected by haemolysis were taken from adult women of reproductive age. No sex or age information was included with either of the compared studies, although it is likely these samples included specimens from men and women. To account for the potential bias introduced using data from only female and all reproductively aged volunteers, we ensured all miRNAs included here have previously been identified in multiple tissues in both male and female samples and are not affected by developmental stage. Investigation using a cohort of mixed age and sex is warranted and may help to further determine which miRNAs are affected by haemolysis.

Interestingly, all signature miRNAs, with the exception of miR-325-5p, have previously been reported as prognostically valuable plasma or serum biomarkers. In this small sampling of recent miRNA biomarker research, we identified several instances where more than one of our haemolysis signature miRNA were identified as disease biomarkers for the same condition in the same experiment (27–29) which, given our findings, and those of previous haemolysis research, further call into question their validity as biomarkers of disease or condition. In conjunction with our research, we found many miRNAs as suggested circulating biomarkers for multiple disease states. For example, miR-122 was given biomarker potential in liver disease, lung cancer and myasthenia gravis (14,27,30), and miR-660 was given biomarker potential in Alzheimer’s disease, breast cancer and lung cancer (28,31,32), respectively. These miRNAs may represent effective biomarkers, but they may simply highlight RBC contamination or be indicative of a general state of inflammation.

Data contained in this study were obtained from two cohorts of female volunteers of reproductive age. Whilst we are working in a relatively narrow experimental domain, we have generalised this method such that removal (from the signature miRNA set) of domain specific miRNA is built in, providing a framework that allows use within research conducted in any human plasma context. Our results highlight that ignoring the issue of miRNA from RBCs leaves researchers open to the risk that newly discovered miRNA disease biomarkers could in fact be biomarkers of haemolysis. In future research, a repeat experiment with samples taken from male and female individuals across a wider age range would expand and strengthen our understanding of the impact of haemolysis on biomarker discovery. Our research both recommends and enables tests for haemolysis to become standard pre-analytical practice.

## Methods

### Sample collection

Peripheral blood (9 mL) was collected with informed, written consent from women undergoing elective terminations of otherwise healthy pregnancies. Blood was collected into standard EDTA blood tubes pre-termination and stored on ice until processed. Whole blood underwent centrifugation at 800 x g for 15 minutes at 4°C before plasma removal and then spun for a further 15 minutes to ensure any remaining cellular debris, including cell membranes from lysed red blood cells, was removed. All samples were stored at -80°C until further processing. Termination samples were collected from the Pregnancy Advisory Centre (PAC), Woodville, South Australia. Blood was also collected with informed, written consent from non-pregnant volunteers at the Adelaide Medical School. Following collection, blood tubes were stored on ice until processing. Whole blood underwent centrifugation at 1015 x g for 10 minutes at 4°C. Approximately 4-6 mL plasma was collected in 2 mL aliquots. 500 μL plasma (the supernatant) were aliquoted into clean tubes and the pellet containing any remaining blood cells at the bottom of the tube was discarded. All samples were stored at -80°C until further processing.

### RNA extraction and library preparation

MicroRNA was isolated from 200 μL plasma samples using the QIAGEN miRNA serum/plasma kit (Qiagen, Hilden, Germany) according to the manufacturer’s instructions and stored at -80°C.

Library preparation and sequencing was performed by Qiagen (Valencia, CA) using the QIAseq miRNA Library Kit and QIAseq miRNA 48 Index IL kits as per manufacturer’s instructions. Amplified cDNA libraries underwent single-end sequencing by synthesis (Illumina v1.9).

### Haemolysis Detection by RT-qPCR

Plasma samples were examined for haemolysis based on the expression levels of two miRNAs: miR-451a and miR-23a. miR-451a (previously named miR-451) is known to be highly expressed in red blood cells, whereas miR-23a is known to maintain stable abundance levels in plasma. After RNA extraction and cDNA synthesis, the delta quantification cycle (Cq) values for miR-23a-miR-451a were calculated independently for each sample. The evaluation of expression levels was performed based on raw Cq values. According to the Qiagen protocol for haemolysis detection (Qiagen, Hilden, Germany) using the ΔΔCq method, samples with a ΔCq <7 for these two miRNAs were considered as clear of contamination; a ΔCq >7 was considered contaminated; a ΔCq =7 was considered borderline.

### miRNA annotation and abundance

Read quality control metrics were assessed using FastQC (33) (http://www.bioinformatics.babraham.ac.uk/projects/fastqc/) to check for per base sequence quality, sequence length distribution and duplication levels. Adapter detection and trimming were performed using Atropos (34). Alignment performed using BWA version 0.7.17-r1188 (GRCh38) (21). Umi_tools was used to collapse duplicate reads mapped to the same genomic location with the same UMI barcode. Quality control metrics were reported using multiQC (35). Read counts for mature miRNAs were determined using an in-house script (36) with microRNA annotation from miRBase, version 22.0 (22,23) (http://www.mirbase.org).

### Analysis of potential confounding factors

All profile and expression analyses were conducted in the R statistical environment (v.4.0.2), using the *edgeR* (v.3.16.5) (37) and *limma* (v.3.30.11) (38) R/Bioconductor packages. Prior to conducting the differential expression analysis between haemolysed and non-haemolysed expression data we considered the effect of participant characteristics such as sex, age, smoking, pregnancy status and ethnicity. Sex was not included here as all samples were taken from female participants. Maternal age was excluded from the final regression model as there was no strong evidence of association with the outcome and hence considering the sample size a simpler model was chosen to preserve degrees of freedom. miRNA identified as differentially expressed between samples from pregnant and non-pregnant women, were removed from the final set of haemolysis signature miRNA. Sequencing batch was also included in all regression models.

### Identification of haemolysis miRNA signature

Prior to defining a collection of haemolysis informative miRNAs, pre-filtering steps were undertaken: 1) mature miRNA with fewer than five reads were reduced to zero independently for each sample, 2) miRNA with fewer than 40 counts per million (CPM) in the haemolysed group (n = 12) were removed from further consideration. This was done to ensure only highly abundant miRNA likely to be present in most samples remain. The Trimmed Means of M values (TMM) normalisation method was used to correct for differences in the underlying distribution of miRNA expression (39). Next, we used *limma* (38) to obtain the fold change of each miRNA between the haemolysed and non-haemolysed groups to identify miRNAs that are more abundant in the plasma affected by haemolysis. To ensure the haemolysis miRNA signature was robust, we took the intersection of the 60 miRNAs from each category of highest expression and lowest adjusted P-value and miRNAs with a log_2_FC > 0.9, revealing a set of twenty high confidence miRNAs. To further refine the set of haemolysis informative miRNAs we used *limma* to calculate the fold change for each miRNA between the samples from pregnant and non-pregnant women not affected by haemolysis and removed any of the high-confidence miRNA which was also differentially abundant in pregnancy. The workflow, source code and input files associated with this research are available at (https://github.com/mxhp75/haemolysis_maternaPlasma.git).

### Classification - Haemolysis Metric

To classify the data coming from samples as haemolysed, borderline or unaffected we first focused on samples from the non-pregnant group. For these, we subset the miRNA read count table into miRNA from the high confidence haemolysis informative miRNA (n=20) and all others (n=169). Using this data partition we calculated the geometric mean of the distribution of read counts using the *psych* package (v1.8.12) (40) and subtracted the geometric mean of the counts of “other” miRNA from that of the “haemolysis informative” miRNA. Next, for samples from the pregnant group, we performed the same calculations described above after first discarding miRNA which were associated with pregnancy.

## Data Availability

All data produced in the present study are available upon reasonable request to the authors and will be made public pending publication.

## Ethics

Ethics approval for the collection of blood from women undergoing elective pregnancy termination between 6–23 weeks’ gestation was provided under HREC/16/TQEH/33, by The Queen Elizabeth Hospital Human Research Ethics Committee (TQEH/LMH/MH). Blood from women forming the general population group was collected after informed consent with ethics approval provided under HREC/H/021/2005, by The University of Adelaide Human Research Ethics Committee.

## Author contribution

CTR created the concept and acquired funding. MS, KP and JB conceived and developed experimental plans. TJK, DMcA and DMcC performed experiments. MS and KP analysed the sequencing data with statistical support from SYL. Manuscript written by MS and KP. All authors read and approved the final version of the manuscript.

## Acknowledgements

We wish to acknowledge the generosity of the women who donated their blood for our research. Without them, this research would not be possible. We also acknowledge valuable input from QIAGEN Genomic Services.

## Supplementary Information Legends

**Supplementary figure 1**: Haemolysis signature feature selection. Raw single-end reads from small RNA-seq libraries are pre-processed using a range of Unix- and python-based computational tools to quantify miRNA expression in each library. Data quality is ensured through quality control steps throughout the workflow. Concurrently with sequencing, ΔCq (miR-23a-miR-451) was assessed by RT-qPCR and incorporated into the differential expression analysis.

**Supplementary Figure 2**: (a) Volcano plot of differential expression. Linear regression identified 138 miRNA which were more highly abundant in haemolysed compared to non-haemolysed samples with FDR < 0.05 (green). (b) MA plot (M (log ratio) and A (mean average)) of Log2 fold change as a function of Log2 average expression indicates most miRNA have an average expression < 10 Log2 CPM. miR-451a and miR-16-5p, both highly red blood cell associated, are highly expressed and more abundant in the haemolysed group (green).

**Supplementary Figure 3:** (a) Volcano plot of differential expression between the pregnant and non-pregnant samples. Linear regression identified 104 miRNA (FDR < 0.05) which were more highly abundant in the pregnant population (red). Haemolysis Metric signature miRNAs are labelled (b) MA plot (M (log ratio) and A (mean average)) of Log_2_ fold change as a function of Log_2_ average expression indicates most miRNA have an average expression < 10 Log_2_ CPM. Unsurprisingly, the most differentially expressed miRNA are miR-517a-3p, miR-517b-3p, miR-516b-5p, miR-518b, which are all members of the highly placenta associated chromosome 19 miRNA cluster.

**Supplementary Table 1**: RT-qPCR Cq data for miR-23a-3p, miR-451a and ΔCq (miR-23a-miR-451).

