## Supplementary material for "Haemolysis detection in microRNA-seq from clinical plasma samples": Supplimentary Figure 1

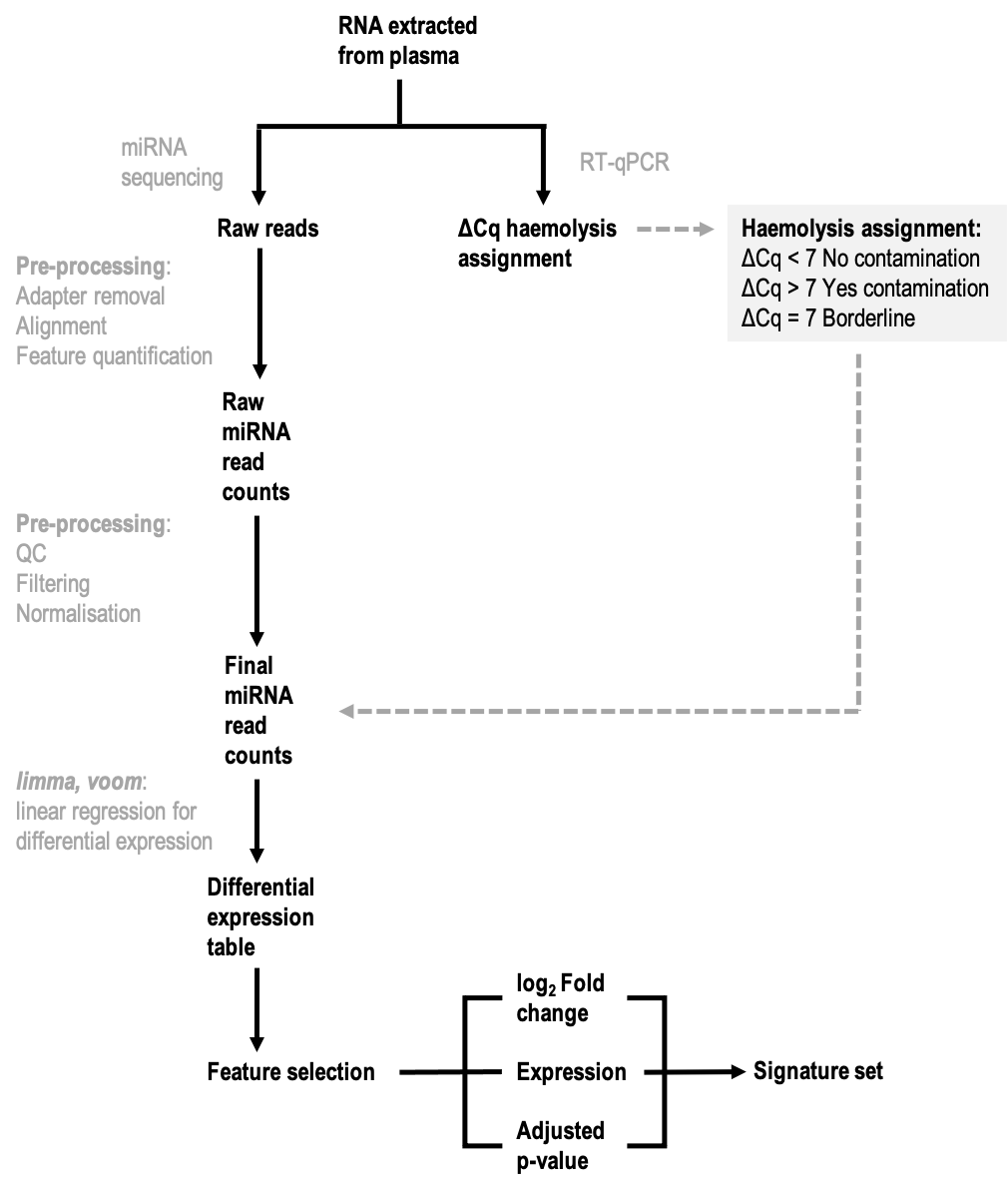

**Supplementary figure 1**: Haemolysis signature feature selection. Raw single-end reads from small RNA-seq libraries are pre-processed using a range of Unix- and python-based computational tools to quantify miRNA expression in each library. Data quality is ensured through quality control steps throughout the workflow. Concurrently with sequencing, ΔCq (miR-23a-miR-451) was assessed by RT-qPCR and incorporated into the differential expression analysis.
