## Supplementary material for "Haemolysis detection in microRNA-seq from clinical plasma samples": Supplimentary Figure 2

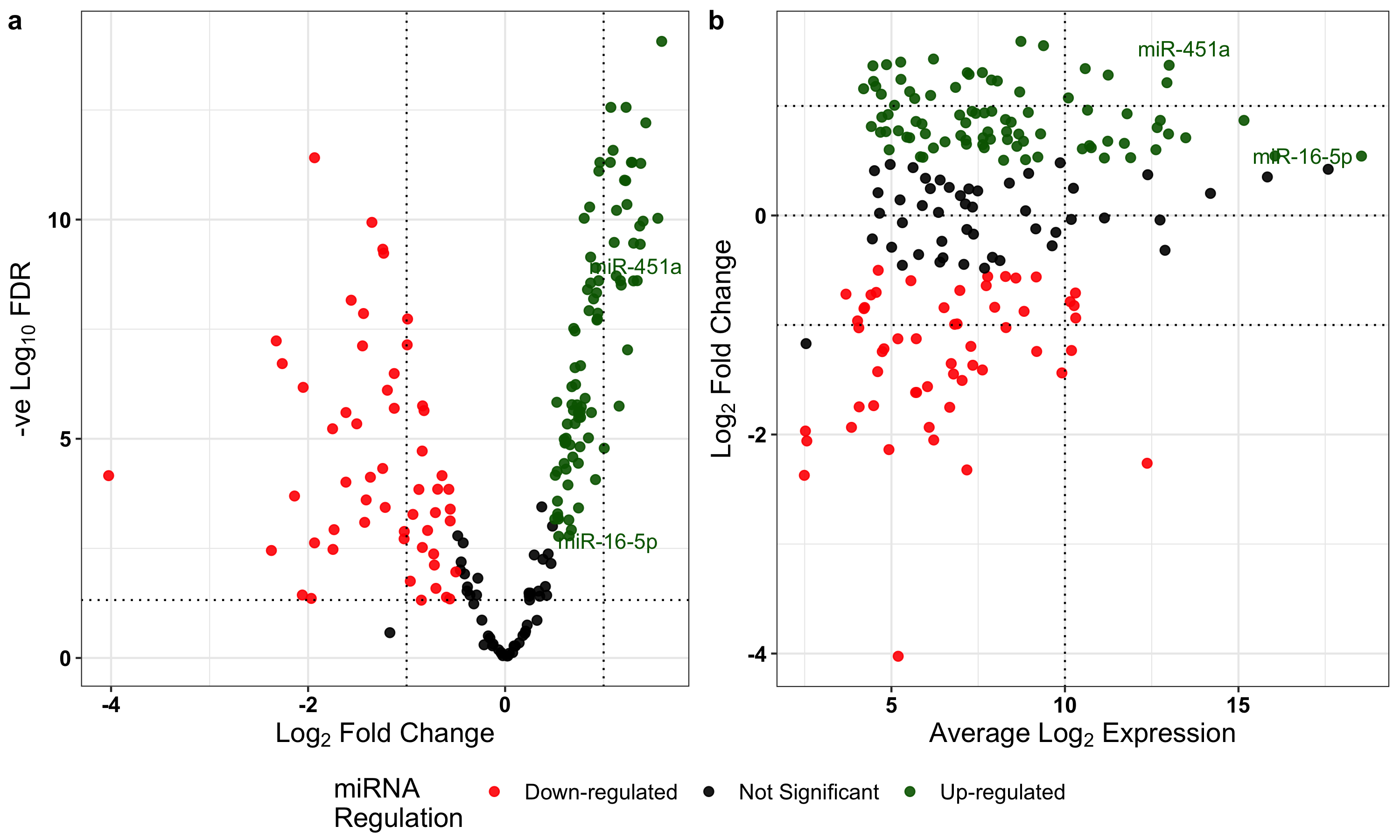

**Supplementary Figure 2:** (a) Volcano plot of differential expression. Linear regression identified 138 miRNA which were more highly abundant in haemolysed compared to non-haemolysed samples with FDR < 0.05 (green). (b) MA plot (M (log ratio) and A (mean average)) of Log_2_ fold change as a function of Log_2_ average expression indicates most miRNA have an average expression < 10 Log_2_ CPM. miR-451a and miR-16-5p, both highly red blood cell associated, are highly expressed and more abundant in the haemolysed group (green).
