## Supplementary material for "Haemolysis detection in microRNA-seq from clinical plasma samples": Supplimentary Figure 3

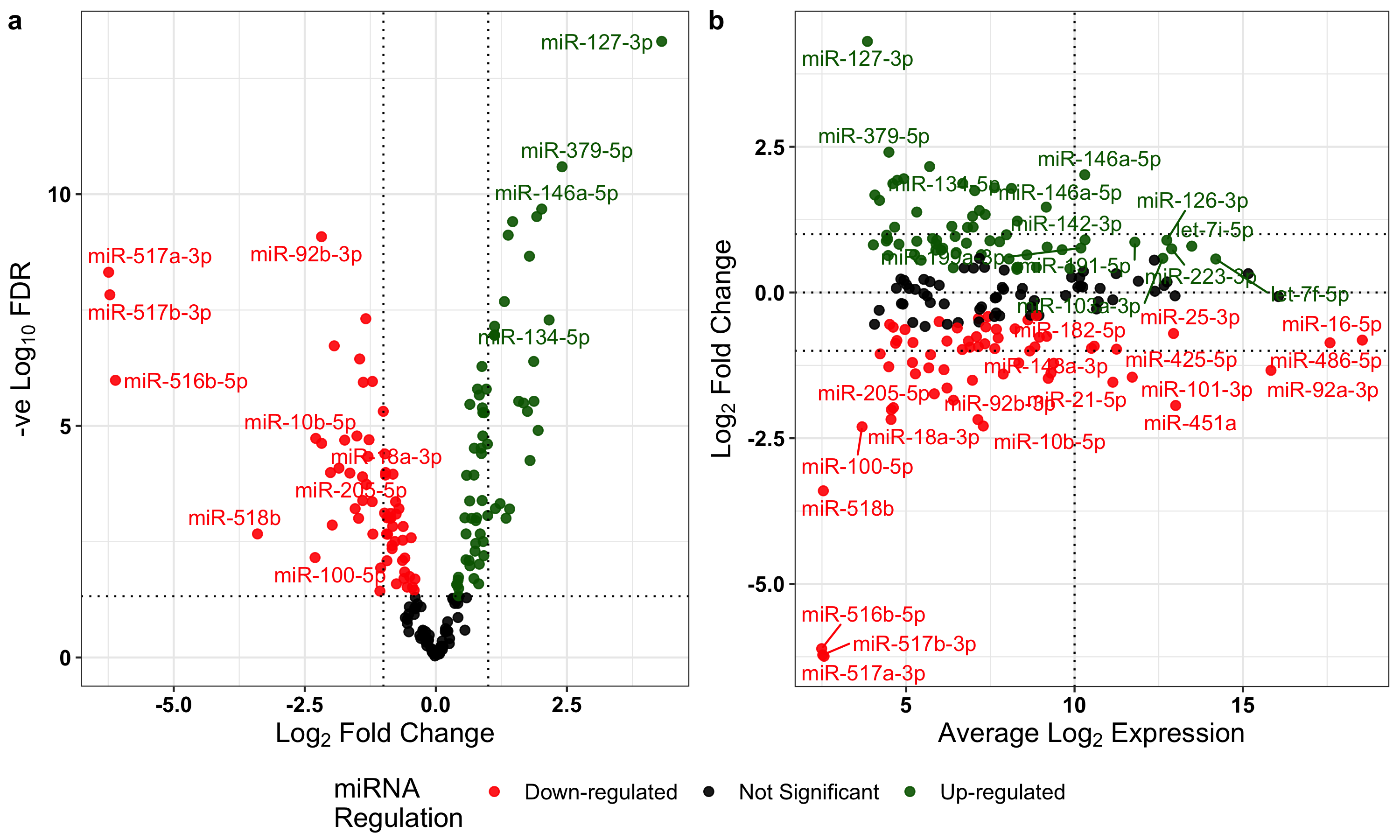

**Supplementary Figure 3**: (a) Volcano plot of differential expression between the pregnant and non-pregnant samples. Linear regression identified 104 miRNA (FDR < 0.05) which were more highly abundant in the pregnant population (red). Haemolysis Metric signature miRNAs are labelled (b) MA plot (M (log ratio) and A (mean average)) of Log_2_ fold change as a function of Log_2_ average expression indicates most miRNA have an average expression < 10 Log_2_ CPM. Unsurprisingly, the most differentially expressed miRNA are miR-517a-3p, miR-517b-3p, miR-516b-5p, miR-518b, which are all members of the highly placenta associated chromosome 19 miRNA cluster.
